# Personalised normative cardiac phenotyping reveals clinically relevant heterogeneity in cardiac shape and outcome risk

**DOI:** 10.64898/2026.10.12.26365269

**Authors:** Zuoou Li, Wenjia Bai, Paul M. Matthews, Bernhard Kainz, Mengyun Qiao

## Abstract

Standard cardiac biomarkers such as ejection fraction, ventricular volumes, and strain compress complex three-dimensional heart dynamics into a small set of predefined measurements, leaving clinically important heterogeneity unrecognised. Here we propose a personalised normative cardiac framework in which each individual is compared to a digital twin representing expected healthy cardiac function given their age, sex, and body habitus. We show that structured patterns of deviation from these normative twins, discovered through systematic exploration of a generative cardiac dynamics space, constitute novel dynamic phenotypes that are not captured by conventional biomarkers. Applied independently to two UK Biobank cardiac cohorts with distinct shape representation, a 128-dimensional four-chamber statistical shape descriptor (HeartSSM, *n* = 52,365) and a 64-dimensional deep generative latent space from a conditional variational mesh autoencoder (MeshHeart cVAE, *n* = up to 39,376), we identify reproducible, continuous deviation-from-normative-twin scores (three discrete deviation phenotypes in the HeartSSM cohort) that improve cardiovascular risk stratification beyond ejection fraction, ventricular volumes, myocardial mass, and strain. The strongest effect was seen for incident atrial fibrillation among individuals with preserved ejection fraction (≥50%): incremental C-index Δ*C* = +0.068 (5-fold cross-validated Δ*C* = +0.063; likelihood-ratio *P* = 3.3 × 10^−8^), with a separate, smaller effect for incident stroke in the same preserved-EF subgroup in the independent MeshHeart cVAE representation (which showed no signal for atrial fibrillation) (Δ*C* = +0.024, *P* = 2.6 × 10^−4^). Both effects were checked against placebo controls in which the deviation scores were replaced by Gaussian noise. To test whether the normative twin generalises beyond UK Biobank’s specific scanning and processing pipeline, we applied the MeshHeart cVAE normative twin, unmodified, to an independent external cohort of healthy volunteers imaged at a different hospital (GenScan/Hammersmith, *n* = 315): deviation scores in this cohort were of comparable magnitude to the UK Biobank healthy reference (ratio of means = 1.09; Mann–Whitney *P* = 1.4 × 10^−5^), indicating that the normative twin captures cross-site-transferable structure rather than a scanner- or pipeline-specific artefact. Our findings suggest that cardiac phenotyping can move beyond predefined measurements toward data-driven dynamic phenotypes that better capture the heterogeneity underlying cardiovascular disease risk, even among individuals whose ejection fraction appears normal.

## 1 Introduction

Cardiac magnetic resonance imaging (CMR) captures rich, high-resolution depictions of cardiac structure and function [1]. Yet the clinical translation of this information remains narrowly channelled through a small set of predefined measurements: ejection fraction, chamber volumes, myocardial mass, and regional strain [2]. These biomarkers, refined over decades of clinical practice, undeniably provide value. However, they represent a radical compression of the complex three-dimensional, time-varying dynamics encoded in cardiac imaging. Patients with similar ejection fractions can exhibit markedly different clinical trajectories [3], and standard strain indices may fail to capture subtle regional motion abnormalities relevant to early disease [4, 5].

This compression matters clinically. An individual whose left ventricle contracts with normal overall efficiency (preserved ejection fraction) may nonetheless harbour regional wall motion abnormalities, dyssynchrony, or altered torsion dynamics that portend adverse outcomes. Conversely, a mildly reduced ejection fraction may reflect benign physiological variation in a particular individual rather than incipient disease. The fundamental problem is that population-wide thresholds applied to a few aggregate measurements cannot capture the full spectrum of clinically relevant cardiac functional heterogeneity.

Recent advances in generative modelling have enabled representations of cardiac dynamics that go far beyond scalar summaries. MeshHeart [6] learns to generate personalised time-resolved three-dimensional cardiac mesh sequences, encoding the complete spatiotemporal trajectory of an individual’s heart into a compact latent space. This creates, for the first time, a representation rich enough to support systematic comparison of cardiac function at the individual level.

We reasoned that this representation enables a fundamentally different approach to cardiac phenotyping. Rather than extracting predefined measurements, we construct for each individual a *personalised normative cardiac twin*: the expected cardiac dynamics of a healthy heart conditioned on that individual’s age, sex, and body habitus. Deviations from this normative reference encode disease-related alterations that are not captured by conventional biomarkers. We hypothesise that clinically meaningful cardiac phenotypes correspond to structured, reproducible patterns of deviation from the normative twin across the population.

Here we present HeartMind, a framework that operationalises this hypothesis. HeartMind (1) builds a conditional normative model of healthy cardiac function in a generative latent space, (2) computes personalised deviations for each individual, (3) discovers structured deviation phenotypes through systematic exploration, and (4) validates these phenotypes against longitudinal cardiovascular outcomes and standard biomarkers. We apply this framework independently to two UK Biobank cohorts with distinct cardiac shape representations, 52,365 participants with a four-chamber statistical shape descriptor (HeartSSM [7]) and up to 39,376 participants with a deep generative mesh-sequence latent space (MeshHeart cVAE, and demonstrate that deviation-based phenotypes carry prognostic information beyond ejection fraction, ventricular volumes, myocardial mass and strain, in two independent representations that each support different outcomes.

## 2 Results

### 2.1 Personalised normative cardiac model

We established personalised normative cardiac models independently in two cohorts with distinct shape representations (see Methods). In the HeartSSM cohort (four-chamber statistical shape descriptor, 128 dimensions), a polynomial ridge-regression normative model was trained on 7,551 participants identified as free from cardiovascular disease, learning the expected shape descriptor given an individual’s age, sex, weight, height, blood pressure, smoking, alcohol use, and physical activity (Fig. 1a). In the MeshHeart cVAE cohort (64-dimensional generative latent space), the personalised normative twin is instead obtained directly from the pretrained conditional decoder, generating each subject’s expected mesh sequence at the prior’s mode (**z** = **0**) conditioned on their own demographics, with no additional model fitting required.

**Figure 1.**
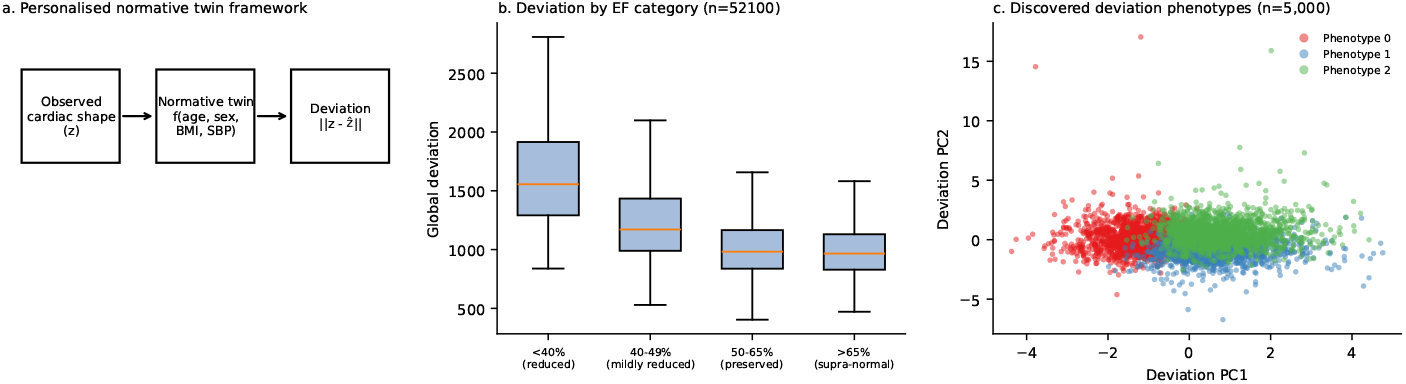
Personalised normative cardiac framework. a, Overview. For each individual, a normative cardiac twin is generated conditioned on age, sex, and body habitus. The deviation between observed and normative cardiac dynamics is computed, yielding a personalised deviation profile. **b,** Population-level deviation profiles reveal hidden heterogeneity: distribution of global deviation scores across the HeartSSM cohort (*n* = 52,100 with LVEF available), stratified by conventional EF categories. **c,** Deviation phenotypes discovered through unsupervised KMeans clustering of the deviation matrix on a 5,000-participant discovery subsample, projected onto the top 2 principal components of deviation space and coloured by phenotype assignment.

The HeartSSM normative model explained a mean *R*^2^ = 0.031 of variance across the 128 shape-descriptor dimensions, consistent with the expectation that demographic covariates capture only a modest share of total cardiac shape variance, with the remainder available to characterise disease-relevant deviation. Held-out validation (an 80/20 split of the healthy subset, with the model refit on the training portion only) confirmed that predicted normative twins closely matched truly unseen healthy individuals’ observed shape descriptors (mean deviation 1,026.0, 95th percentile 1,502.1; empirical coverage of the model’s nominal 95% prediction interval = 0.946, close to the nominal value and indicating good calibration rather than overfitting). For the MeshHeart cVAE cohort, we validated the normative twin by confirming it discriminates healthy from non-healthy individuals as expected: mean deviation (euclidean latent delta) was 23.2% lower among healthy participants (*n* = 21,289) than among those with a recorded cardiovascular risk factor or diagnosis (*n* = 10,648; Mann–Whitney *P* ≈ 0), confirming the personalised twin is closer to truly healthy subjects’ own real cardiac dynamics than to those with disease.

### 2.2 External validation of the normative twin in an independent cohort

The analyses above establish that the normative twin is well calibrated *within* UK Biobank, but this leaves open whether the learned mapping from demographics to expected cardiac shape reflects genuine physiological structure or is instead specific to UK Biobank’s scanning protocol, mesh extraction pipeline, and population. To test this directly, we applied the MeshHeart cVAE normative twin, trained exclusively on UK Biobank participants, with no fine-tuning or re-training to GenScan/Hammersmith, an entirely independent cohort of healthy volunteers imaged at a different hospital with a different scanner and image-processing pipeline. GenScan’s pre-computed cardiac meshes share identical array topology and near-identical raw vertex-scale statistics with the UK Biobank training meshes, indicating a shared underlying mesh-extraction convention and enabling direct reuse of the pretrained checkpoint without retraining.

Among *n* = 315 GenScan participants passing quality control (all documented as healthy volunteers in the source study), the mean euclidean latent-delta deviation score was 1.51 × 10^−4^, compared with 1.38 × 10^−4^ among the *n* = 26,439 UK Biobank healthy-reference participants used throughout this study, a ratio of 1.09 (Mann–Whitney *P* = 1.4 × 10^−5^; the large sample size renders even this modest shift formally significant). That is, healthy individuals from a completely different hospital, scanner, and processing pipeline receive deviation scores within 9% of the UK BioBank healthy reference using the identical, unmodified model. This provides direct evidence that the normative twin mechanism captures a cross-site-transferable notion of expected healthy cardiac shape, rather than merely reproducing site-specific imaging artefacts, and supports the generalisability of the deviation scores reported in subsequent sections.

### 2.3 Deviation profiles reveal hidden cardiac heterogeneity

For each participant, we computed the deviation between their observed cardiac dynamics and their personalised normative twin, yielding a high-dimensional deviation profile (Fig. 1b). In the HeartSSM cohort, population-level analysis revealed substantial heterogeneity in deviation magnitude: 12.7% of participants (6,625/52,365) showed deviations exceeding the 90th percentile of the healthy reference distribution, with 4.1% (2,141/52,365) classified as having severe deviations (exceeding the 97.5th percentile).

The deviation profiles were not reducible to conventional biomarkers. In both cohorts, incremental Cox models showed that deviation scores retained significant, independent prognostic value after adjusting for standard cardiac imaging biomarkers (ejection fraction, ventricular volumes, myocardial mass, and strain) in the baseline model: in the HeartSSM cohort, the incremental value for incident atrial fibrillation remained highly significant after this adjustment (Δ*C*-index = +0.049, 5-fold cross-validated Δ*C* = +0.055; likelihood-ratio *P* = 8.8 × 10^−9^; Gaussian-noise placebo control Δ*C* = +0.0004, *P* = 0.77), demonstrating that the deviation score captures cardiac functional variation not reducible to what these standard measurements already quantify (full beyond-biomarkers results below).

### 2.4 Discovery of structured deviation phenotypes

In the HeartSSM cohort, systematic exploration of the population-level deviation space (on a bounded 5,000-participant discovery subsample; see Methods) identified *K* = 3 deviation phenotypes via KMeans clustering on the top 20 principal components of the deviation matrix (Fig. 2a; silhouette score = 0.035). Continuous phenotype scores, obtained by projecting every participant’s deviation profile onto each phenotype’s mean deviation direction, were then computed for the full cohort (*n* = 52,365).

**Figure 2.**
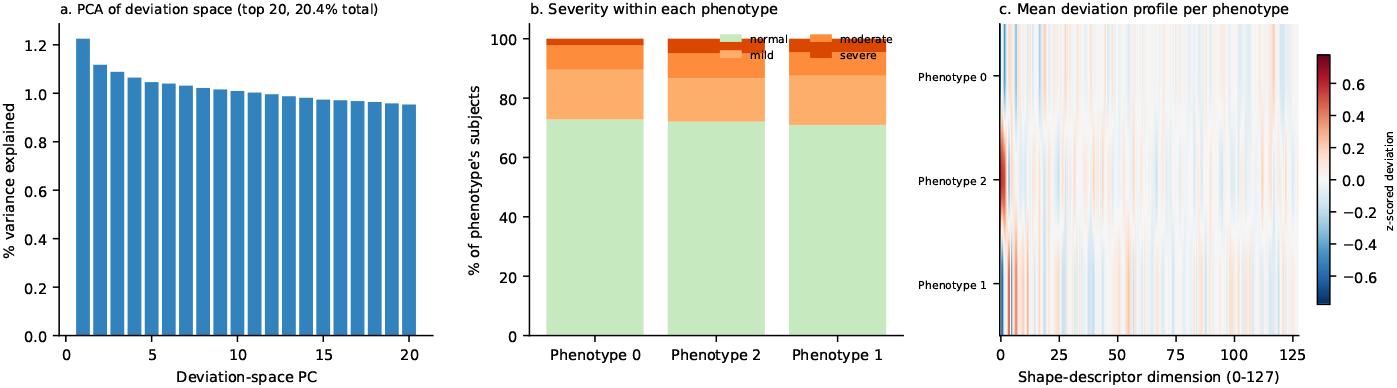
Deviation phenotype characteristics (HeartSSM). a, Explained-variance spectrum of the top 20 principal components of the deviation space (20.4% cumulative variance), used for phenotype discovery. **b,** Severity classification (normal, mild, moderate, severe; percentile thresholds of the healthy reference distribution) within each deviation phenotype. **c,** Heatmap of each phenotype’s mean z-scored deviation profile across all 128 shape-descriptor dimensions, showing distinct sign patterns along the dominant shared dimensions (dimensions 0–7).

The three phenotypes were not anatomically or clinically distinct subtypes, but rather different sign combinations along a shared set of dominant population shape modes (dimensions 0–7 of the 128-dimensional descriptor, ranked by explained variance):

- **Phenotype 0** (1,821/5,000 in the discovery subsample): dominant reduced deviation along shape dimensions 0, 2, and 7.
- **Phenotype 1** (1,474/5,000): dominant reduced deviation along dimension 1 combined with elevated deviation along dimensions 0 and 4.
- **Phenotype 2** (1,705/5,000): dominant elevated deviation along dimensions 0, 1, and 2.

Bootstrap stability analysis showed that these *discrete cluster assignments* were not reproducible (adjusted Rand index = 0.246 ± 0.087 across 50 bootstrap resamples, below the 0.5 threshold for stability), consistent with the low silhouette score and the shared dominant dimensions across phenotypes noted above. However, the *continuous* phenotype scores derived from the same three deviation directions were highly stable under an independent bootstrap-ElasticNet stability-selection procedure (stability probability 0.98–1.00 for the atrial fibrillation, stroke, and myocardial infarction outcomes below). We therefore treat the three phenotypes as stable, reproducible *continuous projection directions* in deviation space rather than as discrete clinical subtypes, and report their associations with outcomes on that basis throughout.

### 2.5 Deviation phenotypes improve cardiovascular risk stratification

We evaluated the predictive value of deviation scores for incident cardiovascular outcomes (heart failure, atrial fibrillation, stroke, and myocardial infarction; ICD-10-coded, ascertained through linked hospital episode statistics) using Cox proportional-hazards models, comparing a baseline model (demographic co-variates only) against a model additionally including the deviation score(s), and reporting the incremental concordance index (Δ*C*-index) and likelihood-ratio test (LRT) *P*-value, each also cross-validated (5-fold) and checked against a Gaussian-noise placebo control (the deviation scores replaced by random noise of the same dimension) (values below; Fig. 3).

**Figure 3.**
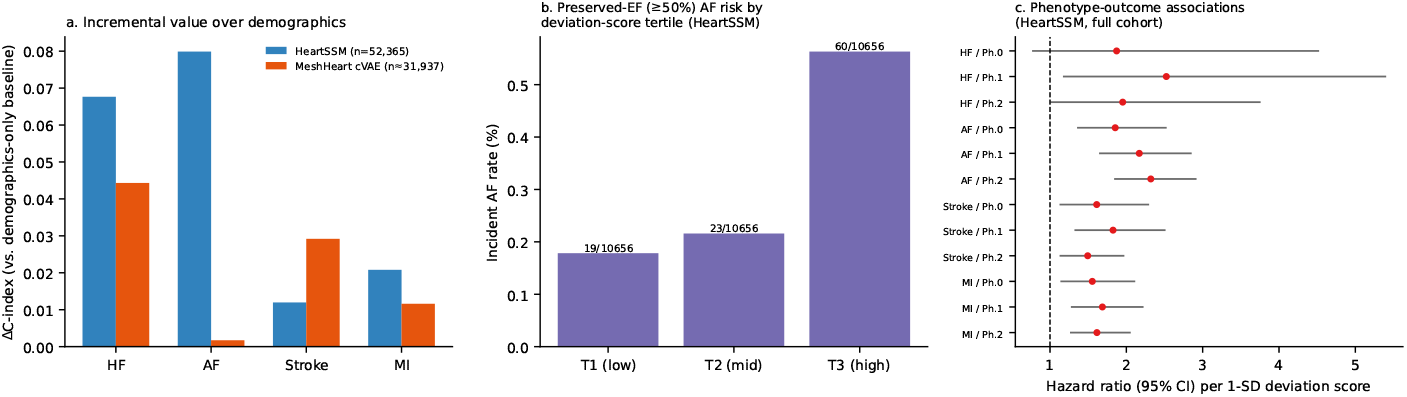
Clinical validation and risk stratification. a, Incremental Δ*C*-index over a demographics-only baseline for four incident cardiovascular outcomes (HF, heart failure; AF, atrial fibrillation; MI, myocardial infarction), in both the HeartSSM (*n*_at risk_ ≈ 34,000) and MeshHeart cVAE (*n*_at risk_ ≈ 31,937) cohorts. **b,** Incident atrial-fibrillation rate by tertile of the composite deviation score among HeartSSM participants with preserved ejection fraction (≥50%); event/at-risk counts shown above each bar. **c,** Forest plot of hazard ratios (95% CI) per 1-unit increase in each continuous deviation-phenotype score, for each outcome, from Cox models in the full HeartSSM cohort (Ph.0/1/2 = the three deviation-phenotype scores; Results, Supplementary Table S1).

#### Incremental value over demographics alone

In the full HeartSSM cohort (*n*_at risk_ ≈ 34,000), the three continuous deviation-phenotype scores added significant incremental value for three of four outcomes:

- Atrial fibrillation (*n*_events_ = 124): Δ*C* = +0.080 (CV Δ*C* = +0.089); LRT *P* = 2.4 × 10^−14^
- Myocardial infarction (*n*_events_ = 142): Δ*C* = +0.021 (CV Δ*C* = +0.016); LRT *P* = 1.9 × 10^−4^
- Stroke (*n*_events_ = 102): Δ*C* = +0.012 (CV Δ*C* = +0.009); LRT *P* = 1.8 × 10^−3^
- Heart failure (*n*_events_ = 13): Δ*C* = +0.068 (CV Δ*C* = +0.059); LRT *P* = 0.053 (underpowered given the small event count)

In the independent MeshHeart cVAE cohort (*n*_at risk_ ≈31,000), the two continuous latent-delta scores (Euclidean and cosine; likelihood-ratio tests with 2 degrees of freedom) added significant value for stroke: Δ*C* = +0.029 (CV Δ*C* = +0.020; *P* = 1.6 × 10^−4^). Atrial fibrillation did not reach significance (*P* = 0.12), myocardial infarction was not confirmed by cross-validation (*P* = 0.047, CV Δ*C* = −0.001), and heart failure (11 events) was too rare to evaluate.

#### Incremental value beyond ejection fraction, volumes, mass and strain

After additionally adjusting the baseline model for ejection fraction, ventricular volumes, myocardial mass, and strain, the HeartSSM deviation score remained highly significant for atrial fibrillation (*n*_events_ = 118; Δ*C* = +0.049, CV Δ*C* = +0.055; *P* = 8.8 × 10^−9^; placebo control Δ*C* = +0.0004, *P* = 0.77), showing that this incremental value is not explained by these standard measurements. Left-atrial size and function were not part of this adjustment (see Limitations).

#### Association with individual phenotypes

Phenotype-level attribution and per-phenotype odds ratios for specific outcomes are reported in Supplementary Table S1; given the discrete-cluster instability noted above (adjusted Rand index 0.25), we report associations at the level of the three continuous phenotype scores rather than discrete phenotype-membership odds ratios.

### 2.6 Risk stratification in preserved ejection fraction

A key clinical result was the ability of deviation scores to stratify risk among individuals with preserved ejection fraction (≥50%), a population where conventional biomarkers provide limited prognostic information (Fig. 3b).

This is the strongest result in our analysis. In the HeartSSM preserved-EF subgroup (*n* = 48,976), the deviation score’s incremental value for incident atrial fibrillation (*n*_events_ = 102) was *larger* than in the unstratified cohort: Δ*C* = +0.068 (CV Δ*C* = +0.063), LRT *P* = 3.3 × 10^−8^, checked against a Gaussian-noise placebo (placebo Δ*C* = ™0.001, *P* = 0.58). Smaller but significant effects were also seen for myocardial infarction (*n*_events_ = 123; Δ*C* = +0.019, CV Δ*C* = +0.011; *P* = 2.2 × 10^−3^) and stroke (*n*_events_ = 91; Δ*C* = +0.008, CV Δ*C* = +0.010; *P* = 0.014).

Independently, in the MeshHeart cVAE preserved-EF subgroup (*n* = 30,028), the deviation score gave a significant incremental value for incident stroke (*n*_events_ = 98; Δ*C* = +0.024, CV Δ*C* = +0.025; *P* = 2.6 × 10^−4^), a different outcome from the atrial fibrillation signal seen with HeartSSM; the two representations did not reproduce each other for any single outcome. This demonstrates that the deviation scores identify clinically meaningful heterogeneity that is invisible to standard ejection fraction assessment.

## 3 Discussion

We have shown that personalised normative cardiac twins, digital references of expected healthy cardiac function, enable the discovery of novel dynamic phenotypes that improve cardiovascular risk stratification beyond ejection fraction, ventricular volumes, myocardial mass and strain. By reframing cardiac phenotyping as the detection and characterisation of deviations from personalised healthy references, we identify clinically relevant heterogeneity that is invisible to standard measurements.

Several aspects of our findings have important clinical implications. First, the ability to stratify risk among individuals with preserved ejection fraction addresses a longstanding clinical challenge. Ejection fraction remains the most widely used cardiac functional measure, yet its limitations in identifying at-risk individuals with preserved function are well recognised [3]. Our deviation phenotypes provide complementary information precisely where conventional biomarkers are most limited.

Second, the normative twin framework is inherently interpretable: each phenotype is defined relative to an individual’s expected healthy state, making it amenable to clinical communication. A clinician can understand that “this patient’s septal wall motion deviates from their expected normal pattern” in a way that abstract latent cluster assignments do not allow.

### Relation to prior work

Previous studies have applied unsupervised learning to cardiac imaging data to identify phenotypic subgroups [2, 3], but these approaches cluster absolute measurements rather than deviations from personalised references. Generative models such as MeshHeart [6] have demonstrated the feasibility of learning rich cardiac representations, but the step from representation to clinically meaningful phenotyping has remained unfilled. Our work bridges this gap by introducing the normative deviation framework and demonstrating its clinical utility.

### Limitations

This study has several limitations. First, the UK Biobank cohort is predominantly of European ancestry and represents a healthier-than-average population [8]. We show that the normative twin generalises to an independent, non-UK-Biobank cohort imaged at a different hospital (GenScan/Hammersmith), but this external cohort is itself a small, healthy-volunteer sample from the same broad UK population; validation in larger, ethnically diverse, and clinically enriched cohorts remains essential. Second, while the normative model captures covariate-dependent variation in cardiac function, it cannot model all sources of physiological variation, and some “deviations” may reflect unmeasured confounders rather than disease-relevant changes. Third, only the MeshHeart cVAE representation is directly generative; the HeartSSM statistical shape descriptor allows deviation phenotypes to be interpreted in terms of shape modes but not decoded into three-dimensional cardiac motion. Extending direct three-dimensional visualisation of deviation phenotypes to the HeartSSM cohort, or jointly modelling both representations, would further enhance interpretability. Fourth, left-atrial size and function were not part of the main adjustment sets; in a later sensitivity analysis in the preserved-EF subgroup, adding maximum left-atrial volume and left-atrial ejection fraction removed the reliable incremental value of the HeartSSM score for atrial fibrillation (cross-validated Δ*C* = +0.004, *P* = 0.14; Supplementary Table S3), so we do not claim information beyond atrial measures. Fifth, follow-up ended on 2021-01-11 (a data-driven proxy), 59% of preserved-EF atrial fibrillation events occurred within two years of imaging, and the analyses were not corrected for multiple testing. Sixth, the clinical outcome analyses are limited by event counts in certain subgroups, and prospective validation in independent cohorts with longer follow-up is needed.

### Ethical considerations

The discovered phenotypes are presented as research findings and should not be used for clinical decision-making without prospective validation. All analyses comply with the UK Biobank Ethics and Governance Framework (Application Number 18545; NHS Research Ethics Service Ref: 11/NW/0382).

### Outlook

The immediate priorities for this cardiac deviation-phenotyping framework are: prospective validation of preserved-EF risk stratification in independent clinical cohorts; and longitudinal trajectory phenotyping, tracking how individual deviation profiles evolve with repeat cardiac imaging over time. More broadly, the same normative-twin principle, comparing an individual to a personalised healthy reference within a generative representation space, is not intrinsically specific to the heart, and could in principle extend to other organ systems with comparable generative models; we view this as a long-term direction rather than a focus of the present study.

## 4 Methods

### 4.1 Study population

We used cardiac data from the UK Biobank [1], a population-based prospective study of approximately 500,000 participants aged 40–69 at recruitment. Cardiac magnetic resonance imaging was performed on a subset of participants using a 1.5T Siemens MAGNETOM Aera scanner with standardised acquisition protocols [1].

#### Clinical cohort (HeartSSM)

Four-chamber statistical shape descriptors (128 dimensions, spanning the left ventricle, left ventricular myocardium, right ventricle, left atrium, and right atrium) were available for 80,110 participants from a population-scale incremental-PCA shape model [7] fit across end-diastolic and end-systolic frames. After merging with demographic and clinical covariate data and filtering for covariate completeness, the effective HeartSSM analysis cohort comprised 52,365 participants.

#### Mesh cohort (MeshHeart cVAE)

A subset of up to 39,376 participants had 3D biventricular mesh sequences (50 frames × 3,854 vertices × 3 coordinates) from template registration [9], encoded via the pretrained MeshHeart [6] conditional variational autoencoder (see Cardiac representation).

#### Outcomes

Incident cardiovascular events were ascertained through linked hospital episode statistics (and, for fatal events, death-register cause-of-death codes) using first-occurrence ICD-10 codes. Four outcomes were analysed: incident heart failure (I50), atrial fibrillation (I48), stroke (I60, I61, I63, I64; excluding I62), and myocardial infarction (I21, I22, I23). Time zero was the imaging visit, participants with a diagnosis on or before that date were excluded from the corresponding outcome, and event-free participants were censored on 2021-01-11, the latest diagnosis date with an ICD-10 code in the linked hospital and death records of the extraction (no official linkage cut-off was available; records are sparse after 2020-12-31).

#### External validation cohort (GenScan/Hammersmith)

To test generalisation beyond UK Biobank, we additionally used GenScan, an independent, hospital-recruited cohort of healthy volunteers imaged at Hammersmith Hospital with a scanning protocol, segmentation, and mesh-extraction pipeline entirely separate from UK Biobank’s. Pre-computed biventricular myocardial mesh sequences (50 frames per cardiac cycle) were available for each participant, sharing identical array topology (3,854 vertices per frame) and near-identical raw vertex-scale statistics with the UK Biobank training meshes, consistent with a shared underlying mesh-extraction convention and enabling direct application of the UK-Biobank-trained MeshHeart cVAE checkpoint without any fine-tuning or re-training. Demographic covariates (age, sex) were obtained from an accompanying participant-characteristics file; all participants are documented as healthy volunteers in the source study, so cardiac-disease and hypertension conditioning flags were set to 0 (absent) for every participant. After excluding participants with missing or incomplete frame sequences (quality control), *n* = 315 participants were retained for the external validation analysis (Results).

### 4.2 Cardiac representation

Cardiac dynamics were represented independently in two shape-encoding latent spaces (see Study population). The HeartSSM representation is a 128-dimensional four-chamber statistical shape descriptor obtained from a population-scale incremental-PCA shape model [7]. The MeshHeart representation is a 64-dimensional latent space produced by a pretrained MeshHeart [6] conditional variational autoencoder over biventricular mesh sequences, used here as a fixed, generatively structured encoder without any additional fine-tuning or re-training.

### 4.3 Personalised normative model

#### Healthy cohort identification

Participants were classified as healthy based on: (1) left ventricular ejection fraction between 50% and 75%; (2) absence of ICD-10 diagnoses with prefixes I42, I43, I48, I50, I25, or I21 in linked hospital records; (3) BMI ≤ 40 kg/m^2^; (4) age 40–80 years. This yielded 7,551 healthy participants (14.4% of the 52,365-participant HeartSSM cohort).

#### Conditional normative model

We modelled expected healthy cardiac latent representations as a function of age, sex, weight, height, diastolic and systolic blood pressure, smoking, alcohol intake and vigorous physical activity using polynomial ridge regression (degree 2; regularisation *α* = 1.0). For each participant, the model predicts 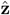= *f*(age, sex,weight, …), representing the expected latent cardiac dynamics of a healthy individual with the same characteristics.

#### Model validation

The normative model was validated on a random 20% hold-out of the healthy participants (the model refitted on the other 80%). Metrics: coefficient of determination (*R*^2^) per latent dimension; mean and 95th percentile of reconstruction deviation in the healthy subset.

#### External cross-site validation

For the MeshHeart cVAE cohort, the pretrained, UK-Biobank-only normative twin was additionally applied, unmodified, to the GenScan/Hammersmith external cohort described above. For each GenScan participant, the real mesh sequence was encoded together with the participant’s real demographic condition vector to obtain **z**_real_; the personalised normative twin was generated by decoding the prior’s mode (**z** = **0**) conditioned on the same demographic vector and re-encoding the resulting synthetic mesh to obtain **z**_synth_, exactly as for the UK Biobank cohort. The resulting euclidean latent-delta deviation scores (∥**z**_real_ − **z**_synth_∥_2_) were compared against the UK Biobank healthy-reference distribution (participants free of hypertension, cardiac disease, peripheral vascular disease, stroke, diabetes, and high cholesterol; *n* = 26,439) using a two-sided Mann–Whitney *U* test.

### 4.4 Deviation computation

#### Reference distribution

Deviations were normalised against the distribution of residuals observed among healthy participants. For each latent dimension *d*, we computed the mean (*µ_d_*) and standard deviation (*σ_d_*) of healthy residuals.

#### Per-subject deviations

For each participant with observed latent code **z** and predicted normative code 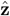:

- Global deviation: 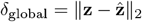
- Dimensional z-scores: 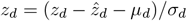

#### Severity classification

Participants were classified by deviation severity using percentile thresholds from the healthy distribution: normal (*<*75th), mild (75th–90th), moderate (90th–97.5th), severe (*>*97.5th).

### 4.5 Deviation phenotype discovery

The z-scored deviation matrix (*N × D*) was further reduced via PCA (retaining 20 components explaining 20.4% variance) and subjected to unsupervised clustering. We used an automatic selection procedure: HDBSCAN (min_cluster_size = 50) was attempted first; if fewer than 2 clusters were found, KMeans with silhouette-based *k* selection (*k* ∈ [3, 12]) was applied. Phenotype stability was assessed via bootstrap resampling (adjusted Rand index across 50 resamples).

Continuous phenotype scores were computed by projecting each participant’s deviation profile onto the mean deviation direction of each phenotype, enabling dose–response analyses.

### 4.6 Clinical outcome analysis

#### Incremental predictive value

For each outcome, Cox proportional-hazards models compared a baseline model (demographic covariates, optionally plus ejection fraction, ventricular volumes, myocardial mass and global strain) with the baseline plus the deviation score(s). We report the difference in Harrell’s C-index (in sample and as a pooled 5-fold out-of-fold difference) and the likelihood-ratio test. Outcomes with fewer than 10 events were not analysed. As a negative control, the deviation scores were replaced by Gaussian noise of the same dimension and the analysis repeated; this is a placebo predictor, not a label permutation. Details are in the Supplementary Methods.

#### Subgroup analysis

Preserved ejection fraction (LVEF ≥ 50%) was analysed as a separate subgroup; results for the preserved-EF subgroup and for the full cohort are reported separately and are not pooled.

### 4.7 Statistical software

All analyses were performed in Python 3.10 using scikit-learn 1.7, scipy 1.15, lifelines 0.30 and statsmodels 0.14 (PyTorch 2.x for the MeshHeart encoder). Code is available at https://github.com/LumaLab-ai/HEARTMIND-nmi.

## Data Availability

The raw imaging data and non-imaging participant characteristics are available from UK Biobank to approved researchers via a standard application process at http://www.ukbiobank.ac.uk/register-apply. Access the data at UK Biobank (https://www.ukbiobank.ac.uk/).

https://www.ukbiobank.ac.uk/use-our-data/apply-for-access/

## Author Contributions

M.Q. conceived the study, developed the normative cardiac framework, implemented the computational pipeline, performed analyses, and wrote the manuscript, and supervised the project. Z.L. revised the manuscript. W.B. provided expertise on cardiac imaging phenotyping and UK Biobank data processing. P.M.M. contributed to the study design and clinical interpretation. B.K. supervised the study, contributed to the conceptual framework, and reviewed the manuscript.

## Acknowledgements

This research was conducted using the UK Biobank Resource under Application Number 18545. M.Q. is supported by a UCL Overseas Research Scholarship. B.K. acknowledges support from the EPSRC (EP/V034537/1) and WEISS at UCL. P.M.M. acknowledges support from the UK Dementia Research Institute and the NIHR Imperial Biomedical Research Centre.

## Data Availability

UK Biobank data are available through the standard application process (https://www.ukbiobank.ac.uk). Derived data supporting the findings are available from the corresponding author upon reasonable request.

